# Self-reported Memory Problems Eight Months after Non-Hospitalized COVID-19 in a Large Cohort

**DOI:** 10.1101/2021.02.25.21252151

**Authors:** Arne Søraas, Ragnhild Bø, Karl Trygve Kalleberg, Merete Ellingjord-Dale, Nils Inge Landrø

## Abstract

**Background:** Neurological manifestations of COVID-19 range from ageusia and anosmia, experienced by most patients, to altered consciousness and rare and severe encephalopathy. A direct affection of the central nervous system (CNS) in the disease has been supported by animal models and MRI findings in patients with mild and severe symptoms. Here we report eight-month data on memory problems for non-hospitalized COVID-19 patients compared to SARS-CoV-2 negative patients and untested volunteers.

**Objective:** To explore the association between non-hospitalized COVID-19 eight months previously and self-reported memory problems.

**Methods:** We followed a cohort of 13156 participants that was invited after (1) being tested for SARS-CoV-2 with a combined oropharyngeal- and nasopharyngeal swab or (2) randomly selected from the Norwegian population (untested). Participants completed online baseline- and follow-up questionnaires detailing underlying medical conditions, demographics, symptoms, and items from the RAND-36 questionnaire on health-related quality of life and known confounders for memory problems.

**Results:** After repeated invitations, the participation rate was 40% (N=794) of SARS-CoV-2 positive, 26% (N=7993) of negative, and 22% (N=4369) of untested randomly selected invitees. All participants completed the baseline questionnaire as a part of inclusion.

The follow-up period was 248 days (SD=18) from baseline, and the follow-up questionnaire was completed by 75% of SARS-CoV-2 positive participants, 65% of negative participants, and 73% of untested randomly selected participants.

At follow-up, 49 (11.5%) of the SARS-CoV-2 positive participants reported memory problems in contrast to 173 (4.1%) of the SARS-CoV-2 negative participants and 65 (2.4%) of the untested randomly selected participants.

In a multivariate model, SARS-CoV-2 positivity remained strongly associated with reporting memory problems at eight months follow-up compared to the SARS-CoV-2 negative group (odds ratio (OR) 4.0, 95% confidence interval (CI) 2.8-5.2) and the untested group (OR 4.9, 95% CI 3.4-7.2).

Compared to the other groups, SARS-CoV-2 positive participants also reported more concentration problems and a significant worsening of health compared to one year ago at follow-up. Feeling depressed, less energy, or pain were reported relatively equally by the different groups.

**Summary:** We find that 11.5% of COVID-19 patients experience memory problems eight months after the disease. SARS-CoV-2 is a new virus, and the long-term consequences of infections are therefore unknown. Our results show that a relatively high proportion of non-hospitalized COVID-19 patients report memory problems eight months after the disease.

## Background

Neurological manifestations of COVID-19 range from ageusia and anosmia, experienced by most patients, to altered consciousness and rare and severe encephalopathy (1). A direct affection of the central nervous system (CNS) in the disease has been supported by animal models and MRI findings in patients with mild and severe symptoms (2-4). Damage to the CNS can lead to long-lasting sequela and a large project on dementia after COVID-19 has been established (5). Many COVID-19 survivors are now at risk for sequelae, and there is an urgent need for a detailed description of the long-term complication (6). Here we report eight-month data on memory problems for non-hospitalized COVID-19 patients compared to SARS-CoV-2 negative patients and untested volunteers.

## Objective

To explore the association between non-hospitalized COVID-19 eight months previously and self-reported memory problems.

## Methods

We followed a cohort of 13156 participants that was invited after (1) being tested for SARS-CoV-2 with a combined oropharyngeal- and nasopharyngeal swab or (2) randomly selected from the Norwegian population (untested). SARS-CoV-2 testing was performed using real-time RT-PCR in four large accredited laboratories in Norway between February 1 and April 15, 2020 (7). Nearly all testing in Norway during the time-period was on symptomatic patients and all adults tested at the four laboratories were invited. Willing participants signed an online electronic consent form. Participants completed online baseline- and follow-up questionnaires detailing underlying medical conditions, demographics, symptoms, and items from the RAND-36 questionnaire on health-related quality of life and known confounders for memory problems. Hospitalized participants are not reported here. The Ethics and ClinicalTrials approval number- and identifiers were REK124170 and NCT04320732.

To determine whether differences between the SARS-CoV-2 positive and negative or untested participants remained after adjustment for confounding, we applied a multivariate logistic regression model that included age, gender, and known confounders for memory problems (RAND-36 items for physical health limitation, pain, feeling energetic, and feeling depressed).

## Results

After repeated invitations, the participation rate was 40% (N=794) of SARS-CoV-2 positive, 26% (N=7993) of negative, and 22% (N=4369) of untested randomly selected invitees. All participants completed the baseline questionnaire as a part of inclusion (Table 1).

**Table 1.**
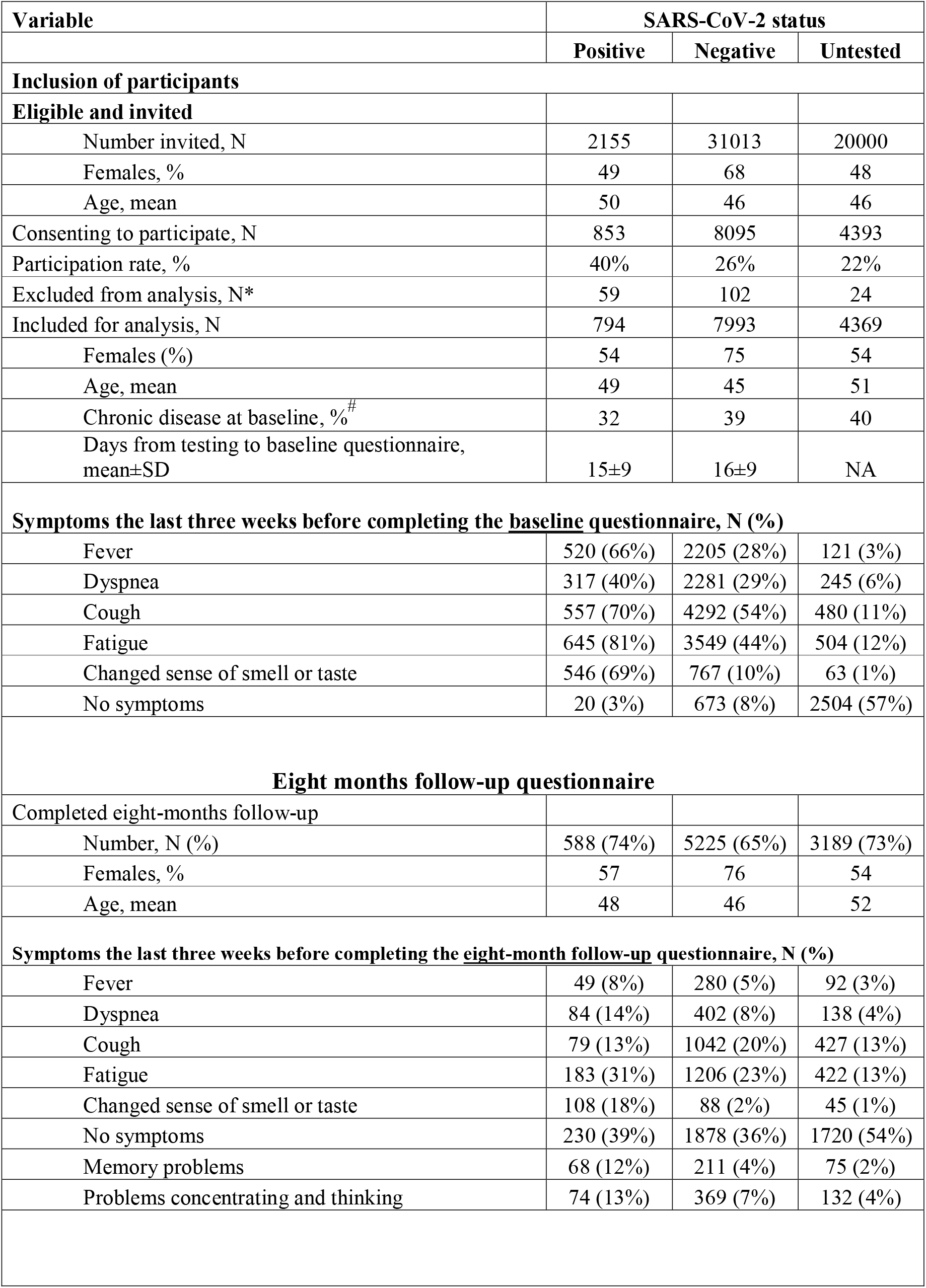
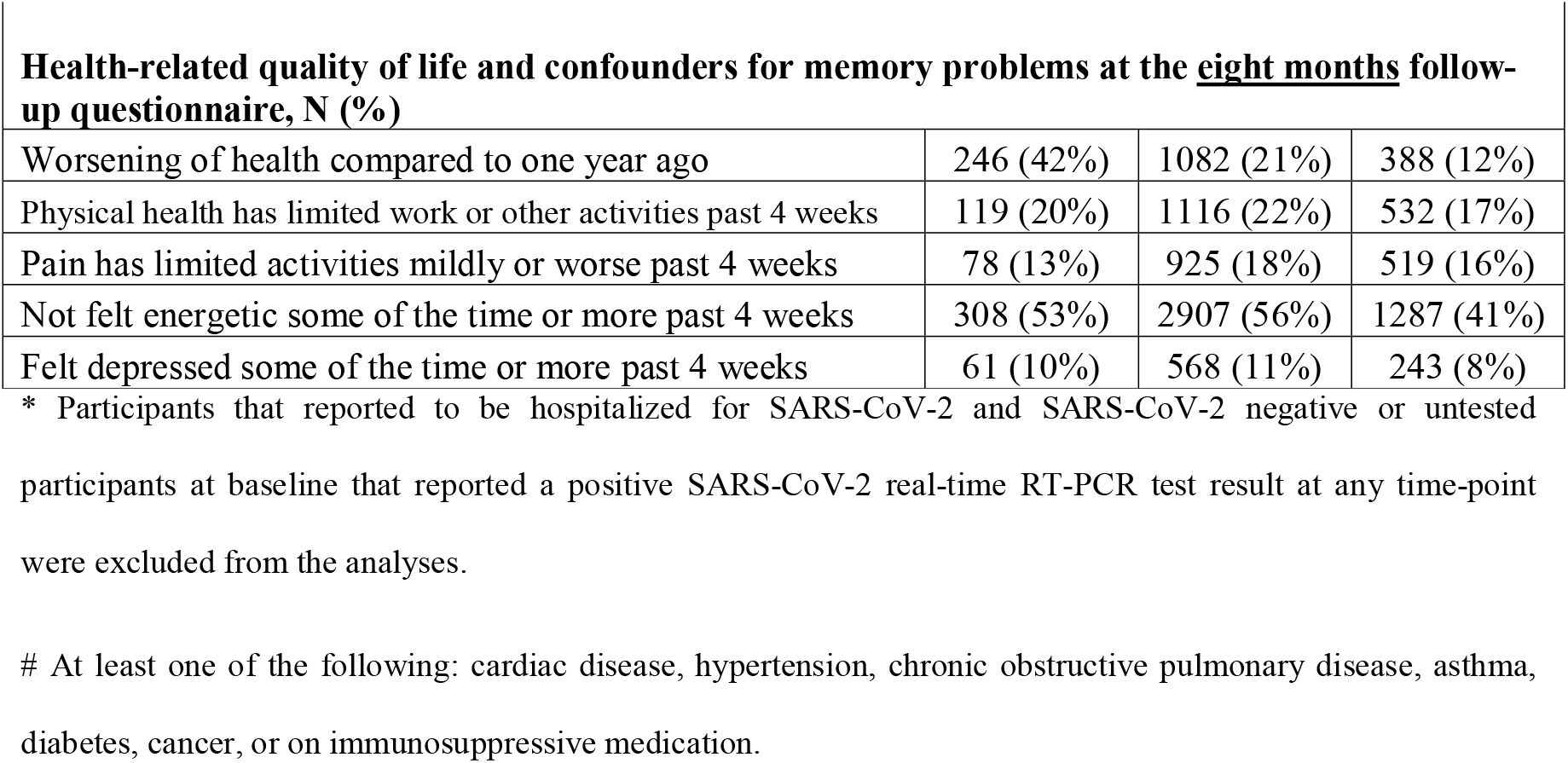
Study population and results. Adult patients having a conclusive SARS-CoV-2 real-time RT-PCR result on a combined oro- and nasopharyngeal swab analyzed in one of four major accredited laboratories and randomly selected untested individuals were invited to complete an online baseline questionnaire from March to May 2020. For the follow-up questionnaire eight months later, five electronic reminders were sent between November 11, 2020, and January 15, 2021. Table legend: SD: Standard deviation, SARS-CoV-2: Severe acute respiratory syndrome coronavirus 2, RT-PCR: Reverse transcription-polymerase chain reaction, N: number, NA: Not applicable

| Variable | SARS-CoV-2 status |  |  |
| --- | --- | --- | --- |
|  | Positive | Negative | Untested |
| **Inclusion of participants** |  |  |  |
| **Eligible and invited** |  |  |  |
| Number invited, N | 2155 | 31013 | 20000 |
| Females, % | 49 | 68 | 48 |
| Age, mean | 50 | 46 | 46 |
| Consenting to participate, N | 853 | 8095 | 4393 |
| Participation rate, % | 40% | 26% | 22% |
| Excluded from analysis, N* | 59 | 102 | 24 |
| Included for analysis, N | 794 | 7993 | 4369 |
| Females (%) | 54 | 75 | 54 |
| Age, mean | 49 | 45 | 51 |
| Chronic disease at baseline, %<sup>#</sup> | 32 | 39 | 40 |
| Days from testing to baseline questionnaire, mean±SD | 15±9 | 16±9 | NA |
| **Symptoms the last three weeks before completing the baseline questionnaire, N (%)** |  |  |  |
| Fever | 520 (66%) | 2205 (28%) | 121 (3%) |
| Dyspnea | 317 (40%) | 2281 (29%) | 245 (6%) |
| Cough | 557 (70%) | 4292 (54%) | 480 (11%) |
| Fatigue | 645 (81%) | 3549 (44%) | 504 (12%) |
| Changed sense of smell or taste | 546 (69%) | 767 (10%) | 63 (1%) |
| No symptoms | 20 (3%) | 673 (8%) | 2504 (57%) |
| **Eight months follow-up questionnaire** |  |  |  |
| Completed eight-months follow-up |  |  |  |
| Number, N (%) | 588 (74%) | 5225 (65%) | 3189 (73%) |
| Females, % | 57 | 76 | 54 |
| Age, mean | 48 | 46 | 52 |
| **Symptoms the last three weeks before completing the eight-month follow-up questionnaire, N (%)** |  |  |  |
| Fever | 49 (8%) | 280 (5%) | 92 (3%) |
| Dyspnea | 84 (14%) | 402 (8%) | 138 (4%) |
| Cough | 79 (13%) | 1042 (20%) | 427 (13%) |
| Fatigue | 183 (31%) | 1206 (23%) | 422 (13%) |
| Changed sense of smell or taste | 108 (18%) | 88 (2%) | 45 (1%) |
| No symptoms | 230 (39%) | 1878 (36%) | 1720 (54%) |
| Memory problems | 68 (12%) | 211 (4%) | 75 (2%) |
| Problems concentrating and thinking | 74 (13%) | 369 (7%) | 132 (4%) |
| **Health-related quality of life and confounders for memory problems at the eight months follow-up questionnaire, N (%)** |  |  |  |
| Worsening of health compared to one year ago | 246 (42%) | 1082 (21%) | 388 (12%) |
| Physical health has limited work or other activities past 4 weeks | 119 (20%) | 1116 (22%) | 532 (17%) |
| Pain has limited activities mildly or worse past 4 weeks | 78 (13%) | 925 (18%) | 519 (16%) |
| Not felt energetic some of the time or more past 4 weeks | 308 (53%) | 2907 (56%) | 1287 (41%) |
| Felt depressed some of the time or more past 4 weeks | 61 (10%) | 568 (11%) | 243 (8%) |

\* Participants that reported to be hospitalized for SARS-CoV-2 and SARS-CoV-2 negative or untested participants at baseline that reported a positive SARS-CoV-2 real-time RT-PCR test result at any time-point were excluded from the analyses.

At follow-up, 49 (11.5%) of the SARS-CoV-2 positive participants reported memory problems in contrast to 173 (4.1%) of the SARS-CoV-2 negative participants and 65 (2.4%) of the untested randomly selected participants (Figure 1).

**Figure 1.**
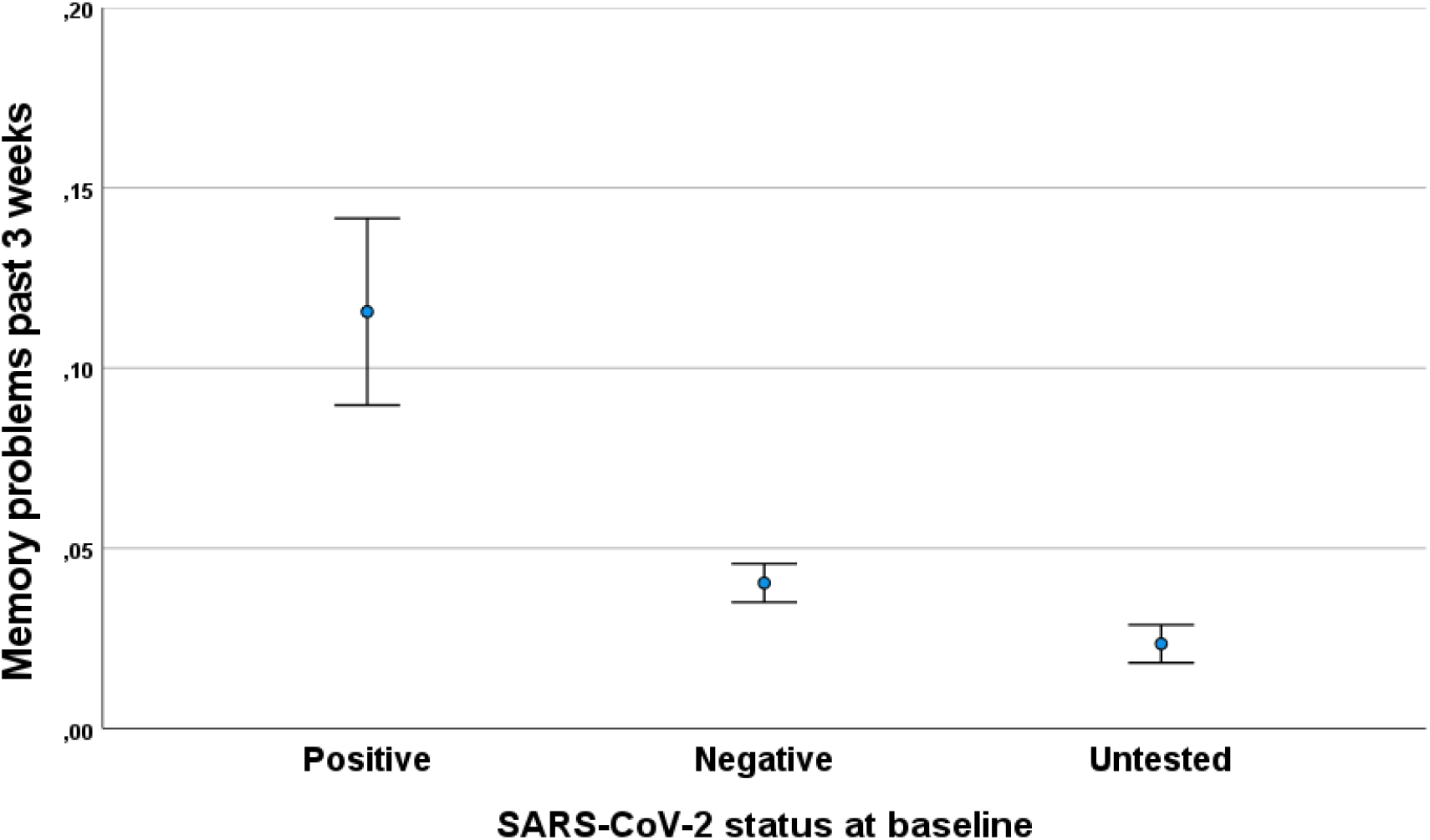
Proportion of participants reporting memory problems eight months after baseline. The figure shows the proportion of participants reporting memory problems during the three weeks before completing the eight-months follow-up questionnaire. Error bars are 95% confidence intervals for the means. SARS-CoV-2: Severe acute respiratory syndrome coronavirus 2

In the multivariate model, SARS-CoV-2 positivity remained strongly associated with reporting memory problems at eight months follow-up compared to the SARS-CoV-2 negative group (odds ratio (OR) 4.0, 95% confidence interval (CI) 2.8-5.2) and the untested group (OR 4.9, 95% CI 3.4-7.2).

Compared to the other groups, SARS-CoV-2 positive participants also reported more concentration problems and a significant worsening of health compared to one year ago at follow-up. Feeling depressed, less energy, or pain were reported relatively equally by the different groups (Table 1).

Patients reporting dyspnea during COVID-19 disease had more memory problems than those not reporting dyspnea (OR 1.5, 95% CI 1.2-1.9).

## Discussion

We find that 11.5% of COVID-19 patients experience memory problems eight months after the disease. Or results are consistent with unpublished surveys without control grops in non-hospitalized and hospitalized patients (8, 9).

Although we ran multivariate models adjusting for several likely confounders, there may still have been unmeasured or residual confounding in the association observed between SARS-CoV-2 status and memory problems at follow-up. An additional limitation of the study is that knowledge of COVID-19 status at baseline could have led to participation or response bias in the follow-up round. A strength is that memory problems have not yet been reported as part of the “long-COVID” syndrome by Norwegian media.

Importantly, memory complaints are known to reflect objective problems and observable changes in everyday function even when controlling for factors associated with self-reported memory problems, such as depression (10, 11).

SARS-CoV-2 is a new virus, and the long-term consequences of infections are therefore unknown. Our results show that a relatively high proportion of non-hospitalized COVID-19 patients report memory problems eight months after the disease.

## Data Availability

Data set: Available from Dr. Soraas (e-mail), data are made available upon agreement of the data provider and within the rules of the General Data Protection Rule and the consent forms.

